# Recent Advancements in Machine Learning-Based Bloodstream Infection Prediction: A Systematic Review and Meta-analysis of Diagnostic Test Accuracy

**DOI:** 10.1101/2024.04.15.24305877

**Authors:** Rajeev Bopche, Jan Kristian Damås

## Abstract

**Purpose:** Bloodstream infections (BSIs) present significant public health challenges. With the advent of machine learning (ML), promising predictive models have been developed. This study evaluates their performance through a systematic review and meta-analysis.

**Methods:** We performed a comprehensive systematic review across multiple databases, including PubMed, IEEE Xplore, ScienceDirect, ACM Digital Library, SpringerLink, Web of Science, Scopus, and Google Scholar. Eligible studies focused on BSIs within any hospital setting, employing ML models as the diagnostic test. We evaluated the risk of bias with the Quality Assessment of Diagnostic Accuracy Studies (QUADAS-2) checklist and assessed the quality of evidence using the Grading of Recommendations, Assessment, Development, and Evaluation (GRADE) approach. Models reporting the area under the receiver operating characteristic curve (AUROC) were included in the meta-analysis to identify key performance drivers.

**Results:** After screening, a total of 30 studies were eligible for synthesis, from which 41 models and 8 data types were extracted. Most of the studies were carried out in the inpatient settings (n=17; 56%), followed by the emergency department (ED) settings (n=7; 23%), and followed by the ICU settings (n=6; 20%). The reported AUROCs in the hospital inpatients settings, ranged from 0.51-0.866, in the ICU settings AUROCs ranged from 0.668-0.970, and in the emergency department (ED) settings the AUROCs of the models ranged from 0.728-0.844. One study reported prospective cohort study, while two prospectively validated their models. In the meta-analysis, laboratory tests, Complete Blood Count/Differential Count (CBC/DC), and ML model type contributed the most to model performance.

**Conclusion:** This systematic review and meta-analysis show that on retrospective data, individual ML models can accurately predict BSIs at different stages of patient trajectory. Although they enable early prediction of BSI, a comprehensive approach to integrate data types and models is necessary. Systematic reporting, externally validated, and clinical implementation studies are needed to establish clinical confidence.

## 1. INTRODUCTION

Bloodstream infections (BSIs) pose a severe public health threat, often escalating to critical conditions such as sepsis and septic shock, particularly if not promptly recognized and treated. The rapid progression of these infections contributes to high morbidity, mortality, and significant healthcare costs, making BSIs a pivotal challenge in clinical care [1, 2]. Currently available clinical decision tools for BSIs and sepsis are primarily based on changes in vital signs and abnormal blood test results, which lack sufficient accuracy [3, 4]. Enhancing prediction capabilities could lead to more efficient resource allocation and cost reduction. Accurate initial stratification helps direct resources to high-risk BSI patients and minimizes unnecessary tests and treatments for those at low risk. Moreover, ambiguous results from blood cultures with contaminants can prolong hospital stays and lead to unnecessary antibiotic administration [5, 6]. Therefore, improving the predictive value of these tests and reducing their unnecessary use are critical for optimizing healthcare resource allocation and minimizing costs.

Despite the extensive application of machine learning (ML) across various healthcare diagnostics, its potential in BSI prediction has not been fully exploited, particularly when compared to its use in predicting sepsis, where it has been more thoroughly researched [7–14]. This gap underscores a critical oversight; early detection of BSIs can significantly limit their progression to sepsis, thus reducing patient suffering and healthcare expenses [16]. This systematic literature review (SLR) and meta-analysis (MA) seek to bridge this gap by highlighting recent advancements in ML models for BSI prediction. Our analysis spans a range of algorithms and examines the influence of different data types used within these models, providing a comprehensive overview of the current landscape and suggesting directions for future research.

### 1.1. Related works

Recent systematic reviews on ML for BSI prediction have illuminated the capabilities of validated models in clinical environments. For example, Eliakim-Raz et al. (2015) emphasize the variability in study populations, model parameters, and validation processes, which highlights the complexity in creating universally effective ML models for BSI prediction [16]. In contrast, the review by Coburn et al. (2012) focuses on the critical clinical and laboratory markers needed to determine the necessity for blood cultures in suspected bacteremia cases, further supporting the role of ML in enhancing diagnostic accuracy and reducing false positives [2]. These studies demonstrate the untapped potential of ML to streamline diagnostic processes and improve patient care in BSI contexts, advocating for its increased integration into BSI prediction strategies.

## 2. METHODS

The guidelines provided by the Preferred Reporting Items for Systematic Reviews and Meta-Analysis of Diagnostic Test Accuracy studies (PRISMA-DTA) statement [15] was followed for the conduct of this literature review and meta-analysis.

### 2.1. Search Strategy

The literature search spanned nine databases to cover a wide array of relevant literature. The databases included: PubMed, IEEE Xplore Digital Library, ScienceDirect, ACM Digital Library, SpringerLink, Web of Science, Scopus, and Google Scholar. These databases were selected to encompass a vast range of medical and computer science literature, including journal articles and conference proceedings. To ensure a comprehensive and relevant literature retrieval, we first identified key terms and their possible alternatives. This approach ensures coverage of a broad spectrum of pertinent studies. The core search terms, along with their alternatives, were: Bloodstream Infection (BSI) – Blood culture, Bacteremia, Blood culture test; Prediction – Diagnosis, Identification, Detection; Machine Learning – Artificial Intelligence, Deep Learning, Supervised Learning, Computational Intelligence; Clinical Data – Electronic Health Records (EHR), Electronic Patient Records (EPR), Electronic Medical Records (EMR). Adjustments were made to the query to accommodate the syntax of different database search engines. The search query was constructed by combining these terms using Boolean operators:

− (Bloodstream Infection OR Blood culture OR Bacteremia) AND (Prediction OR Diagnosis OR Identification OR Detection) AND (Machine Learning OR Artificial Intelligence OR Deep Learning OR Supervised Learning OR Computational Intelligence) AND (Clinical Data OR Electronic Health Records OR Electronic Patient Records OR Electronic Medical Records)

The selection of relevant studies involved a multi-step process. First, The execution of the search query in each database. Then, Initial screening based on titles and abstracts to exclude non-relevant records. In the third step, detailed screening involving full-text reviews was carried out. The reasons for exclusion were documented. Following this, references in selected articles were examined to identify additional relevant studies, implementing snowballing technique for accessing new literature. This structured approach ensured comprehensive coverage of the topic, allowing the retrieval of documents that met the inclusion criteria for data extraction and analysis to address the research questions of this SLR.

### 2.2. Study Selection

#### 2.2.1. Inclusion Criteria

- **Study Focus:** This review specifically targeted studies that utilized machine learning models for the prediction of bacteremia, BSI, or positive blood culture results. The primary objective was the development or validation of these predictive models.

- **Types of Publications**: The review was confined to peer-reviewed journal articles and conference papers.

- **Target Population:** The studies included focused on adult populations.

- **Model Specification:** Included studies clearly described the ML model(s) used, including the type of algorithm, data inputs, and validation methods.

- **Outcome Measures:** Studies reported specific outcomes related to the accuracy, sensitivity, specificity, or predictive value of the ML models for BSI prediction.

#### 2.2.2. Exclusion Criteria

- **Focus on Sepsis and Severe Sepsis:** Studies aimed primarily at predicting the onset of sepsis or severe sepsis, rather than BSI, were excluded. This was to concentrate on predictive research for BSI, an antecedent condition, rather than sepsis, a consequent condition.

- **Pediatric Studies:** Research focusing on neonatal, pediatric, or infant populations was excluded, as the review was limited to adult populations.

- **Post-Diagnosis Prediction:** Studies focusing on predicting outcomes or complications for patients already diagnosed with BSI were excluded. The emphasis was on the initial prediction of BSI.

- **Non-Machine Learning Models:** Research employing statistical or other predictive methodologies without using ML algorithms was not included.

- **Non-Peer Reviewed Literature:** Studies that had not undergone a peer review process, including grey literature, technical reports, and unpublished manuscripts, were excluded.

- **Opinion Pieces and Theoretical Works:** Commentaries, opinion pieces, and purely theoretical works lacking empirical data or validation were omitted.

#### 2.2.3. Timeframe and Language Restrictions

- **Language Restrictions:** The review included only studies published in English. This criterion was applied to ensure the feasibility of thorough analysis and comprehension of the research findings.

- **Timeframe of Publication:** The review focused on studies published within the last five years, capturing the most recent advancements in machine learning applications for bloodstream infection prediction.

### 2.3. Data Extraction

A structured approach was used to extract relevant data from identified studies. The data extraction process involved the following key elements:

− **Study Identification:** Details including authors, year of publication, and title of the study.
− **Population Characteristics**: Information on the patient demographics, clinical settings, and specific population subgroups studied.
− **Dataset Description:** Description of the dataset used in the study, including size, source, prevalence rates, and period of data collection.
− **Algorithms Employed:** Identification of the specific ML algorithms used in the study.
− **Reported Metrics**: Reporting on the performance metrics used in the study.
− **Key Findings and Predictors:** Summary of the main findings, including key predictors identified for BSI prediction.

### 2.4. Quality Assessment

To evaluate risk of bias, we used the Quality Assessment of Diagnostic Accuracy Studies (QUADAS-2) criteria [17]. Domains included patient selection, index test, reference standard, and flow and timing. In line with the recommendations from the QUADAS-2 guidelines, questions per domain were tailored for this paper and can be found in (Supplementary Information). If one of the questions was scored at risk of bias, the domain was scored as high risk of bias. At least one domain at high risk of bias resulted in an overall score of high risk of bias, only one domain scored as unclear risk of bias resulted in an overall score of unclear risk of bias for that paper. We used the Grading of Recommendations Assessment, Development and Evaluation (GRADE) methodology to assess the quality of evidence per hospital setting for all studies reporting the area under the curve of the receiver operating characteristic (AUROC) as their performance metric [18]. In line with the GRADE guidelines for diagnostic test accuracy, we included the study design, domains risk of bias (limitations), and inconsistency of the results. One level of evidence was deducted for each domain with serious concerns or high risk of bias, no factors increased the level of evidence (Supplementary Information). Overall level of evidence is expressed in four categories (high, moderate, low, very low).

### 2.5. Meta-analysis

For meta-analysis we conducted univariate and multivariate random effects model analysis fitted using restricted maximum likelihood estimation (REML) for the contribution of covariates towards model performance [19]. We grouped medical data types utilized to train the models such as (demographics as features, vital signs as features, laboratory tests as features, use of textual data, use of time series data, etc.) and included them as covariates. We performed pooled regression analysis to study the most significant covariates per study settings.

## 3. RESULTS

### 3.1 Study selection

A comprehensive literature search was carried out from November 2023 to March 2024, yielding 348 articles post-duplication removal. Following an in-depth assessment for eligibility, 30 studies conformed to our inclusion criteria, references [20–49]. The selection process, including reasons for exclusion at each stage, is outlined in the PRISMA flow diagram (Figure 1). Notably, the majority (29 out of 30) of the included articles employed retrospective study designs, with only two conducting prospective validations of their models [40, 41]. A single study presented a prospective cohort design [29].

**Fig 1.**
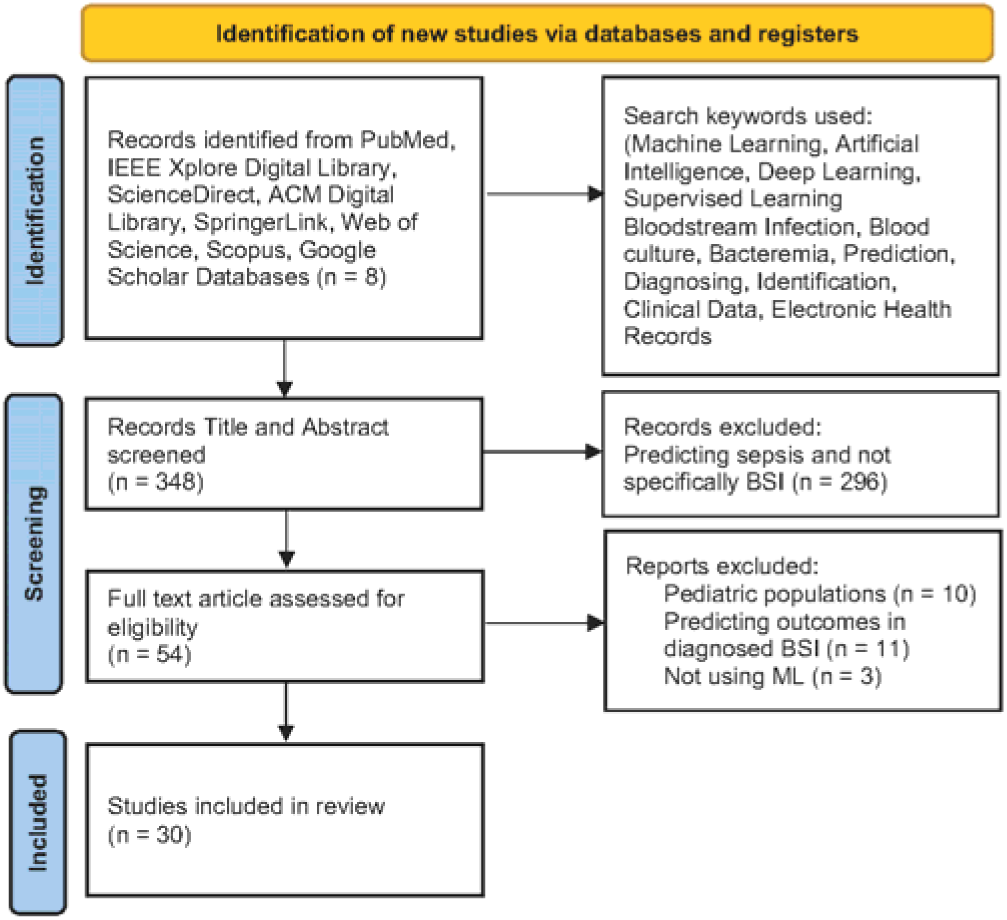
PRISMA flow diagram illustrating the systematic review methodology, depicting the screening, eligibility assessment, and inclusion of studies, alongside exclusion justifications at each phase

### 3.2 Study characteristics

The studies predominantly took place in inpatient settings (56%, n=17), with the emergency department (ED) (23%, n=7) and intensive care units (ICU) (20%, n=6) also represented. Within inpatient settings, nine studies examined general populations [20–28], and others focused on specific patient cohorts including those with central venous catheters (CVC) [30, 31], systemic inflammatory response syndrome (SIRS) [29], and various other conditions. In ED settings, seven studies were identified [37–43], two of which addressed specific patient groups. ICU-focused research numbered six studies [44–49], again with two concentrating on specific patient subsets. Bacteremia was the primary condition investigated in 24 studies, alongside candidemia and central line-associated bloodstream infection (CLABSI) in others. Contaminants were generally considered negative results, except in two studies [23, 45]. Table 1 presents a synthesis of the study characteristics, including target conditions, patient/sample sizes, data sources, and prevalence, categorized by setting.

**Table 1.**
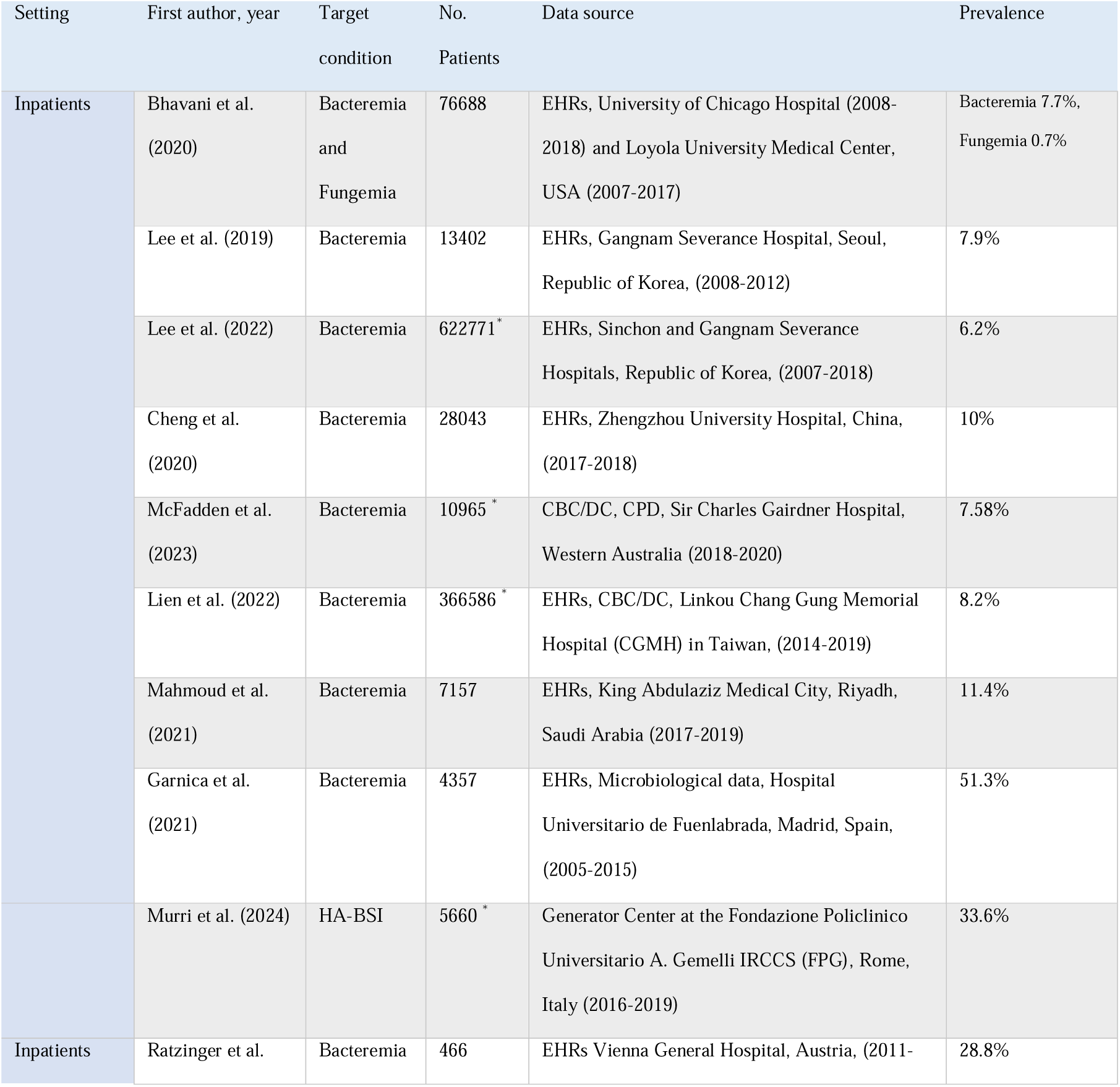
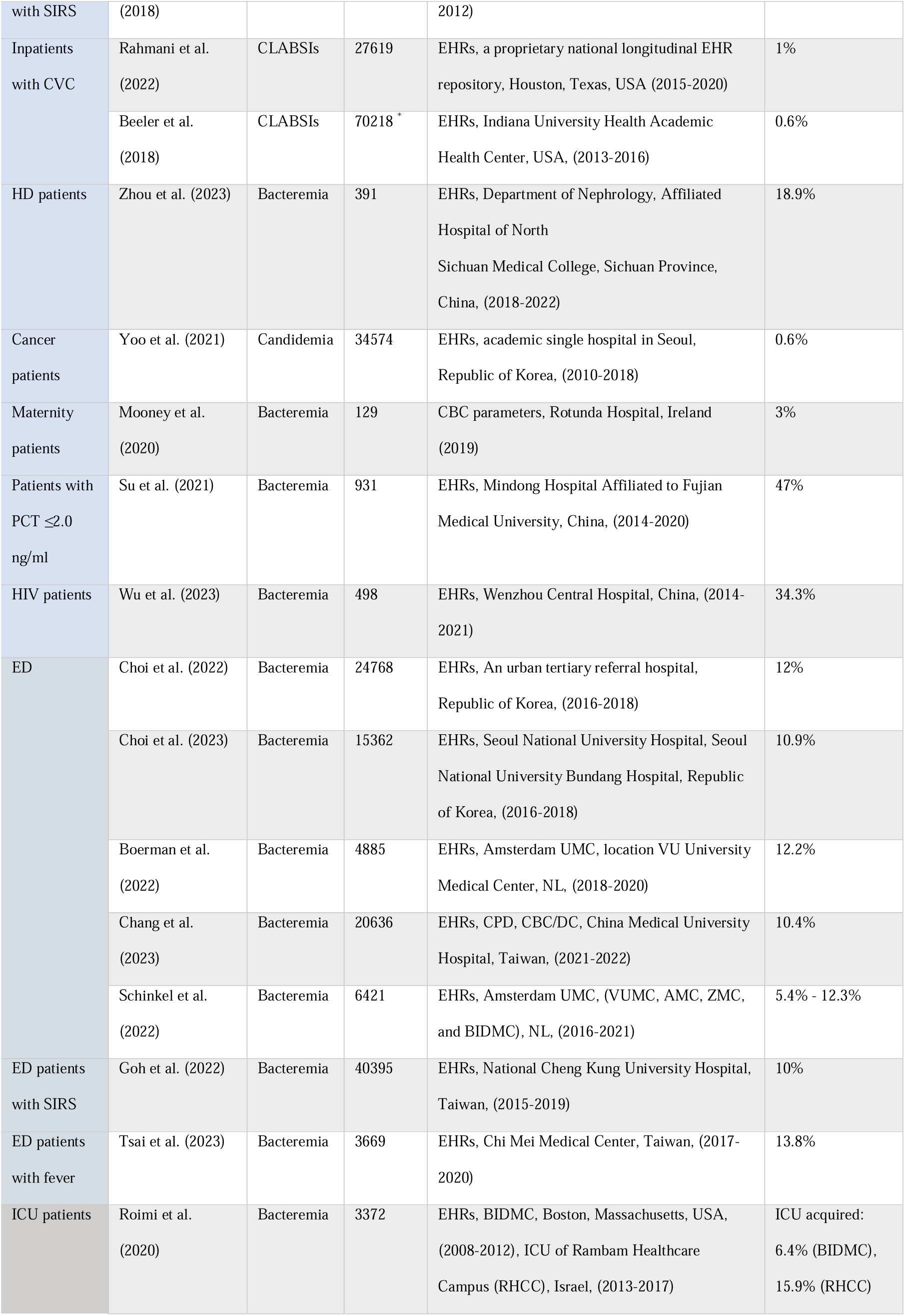
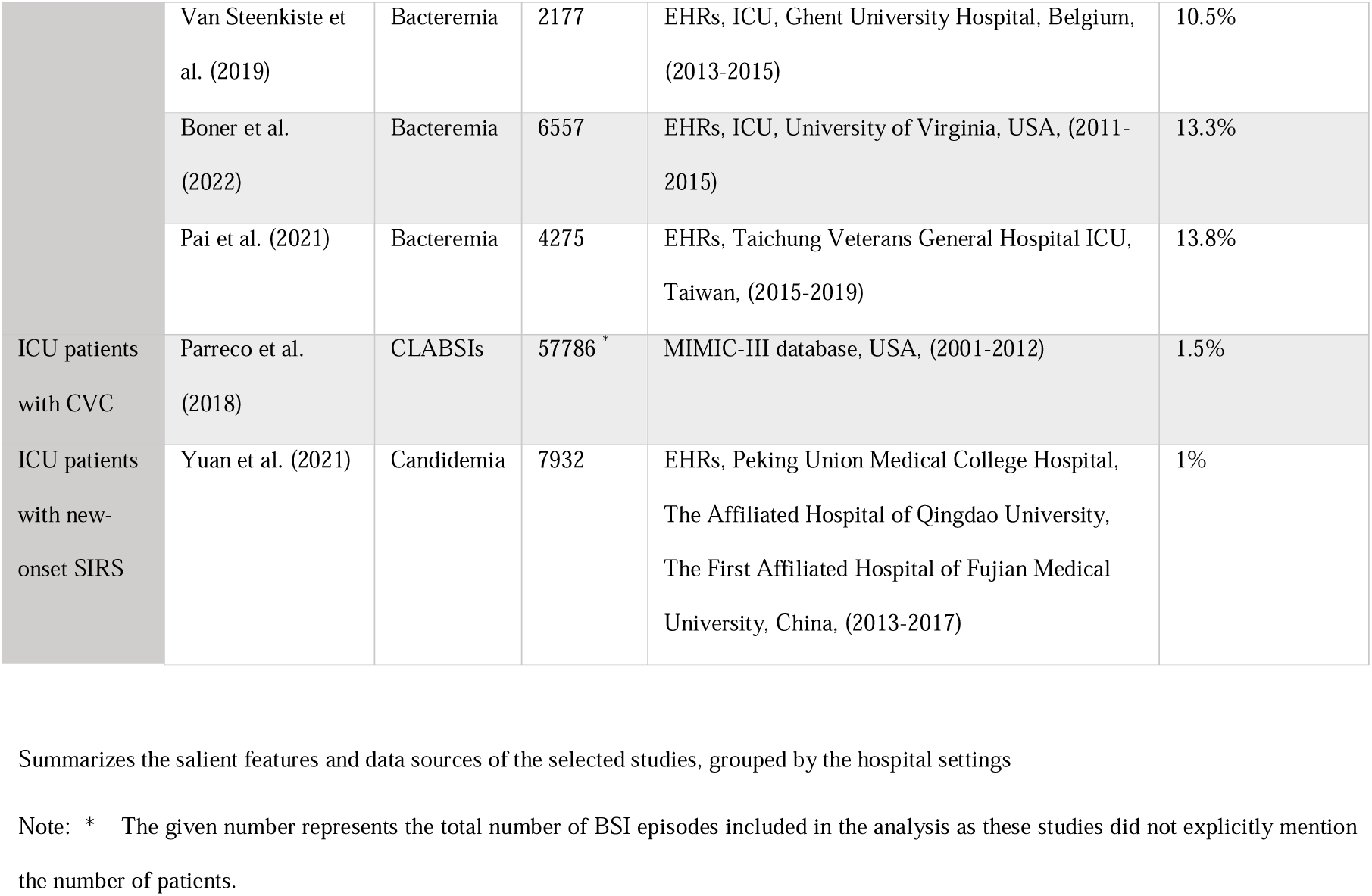
Key characteristics and data sources of selected studies.

Most of the reviewed studies were conducted within a single institution; however, three studies utilized datasets encompassing two hospital systems [20, 22, 44], and two studies expanded their analysis to incorporate multi-center data [41, 49]. External validation, which is critical for the generalizability of findings, was performed in four studies [38, 40, 41, 44]. Compliance with the Transparent Reporting of a multivariable prediction model for individual prognosis or diagnosis (TRIPOD) guidelines, which enhance the reliability of predictive modeling, was confirmed in five studies [20, 28, 37, 39, 41, 50]. Data diversity is key in model training and validation. Within this context, three studies utilized the publicly available Medical Information Mart for Intensive Care (MIMIC) database [41, 44, 48], while four studies included Complete Blood Count/Differential Count (CBC/DC) data [24, 25, 34, 40]. Furthermore, two of these studies also incorporated Cell Population Data (CPD) [24, 40], highlighting the integration of detailed hematologic parameters. Unstructured data utilization was observed in one study that utilized textual data [23]. In terms of data accessibility, most studies employed proprietary hospital data. Data sharing policies varied: one study explicitly stated that their data would not be shared [47], one study offered openly available data [34], and ten studies indicated that deidentified data could be provided upon reasonable request [24, 25, 27, 28, 29, 38, 39, 41, 42, 49], thus contributing to transparency and reproducibility. Notably, six articles did not specify the number of patients included in their analysis [22, 24, 25, 28, 31, 48].

### 3.3. Models and data types

The reported AUROCs in the hospital inpatients settings ranged from 0.51-0.866, while models on select group of participants like HD patients [32], patients with low Procalcitonin (PCT) [35], and HIV patients [36] achieved high AUROCs of 0.91, 0.92, and 0.91 respectively. For the emergency department (ED) settings the AUROCs of the models ranged from 0.728-0.844. For the ICU settings AUROCs ranged from 0.668-0.970. All articles reported high prediction performance (AUROC > 0.7) except for one article [26]. All except two articles [23, 34] reported AUROC as their performance metric. Figure 2 presents a horizontal bar chart that delineates the distribution of AUROC values across the various studies reporting AUROC, with the bar length indicating the range of AUROC values reported across various ML models utilized in the study. The gradient color scheme distinguishes the studies based on the clinical settings, ED (red), inpatient (blue), and ICU (yellow). On the right axis the number of patients, or the number of BC episodes/samples analyzed in the study. Figure 3 depicts the utilization frequency of different ML algorithms categorized into tree-based models, traditional ML algorithms, and neural networks. Notably, tree-based models such as RF and XGB displayed a higher incidence of application. LR was predominantly favored among traditional ML algorithms and within neural networks, ANN and MLP were the commonly employed architectures. Figure 4 presents a horizontal bar graph enumerating the occurrence of various performance metrics in the selected studies. The AUROC was the most reported metric. Figure 5 illustrates the types and diversity of data inputs used with the ML models across the selected studies via a stacked bar chart. The derived risk factors represent the derived clinical features included in the models. The ML algorithms and top predictors for each study grouped by settings are given in Table 2.

**Fig 2.**
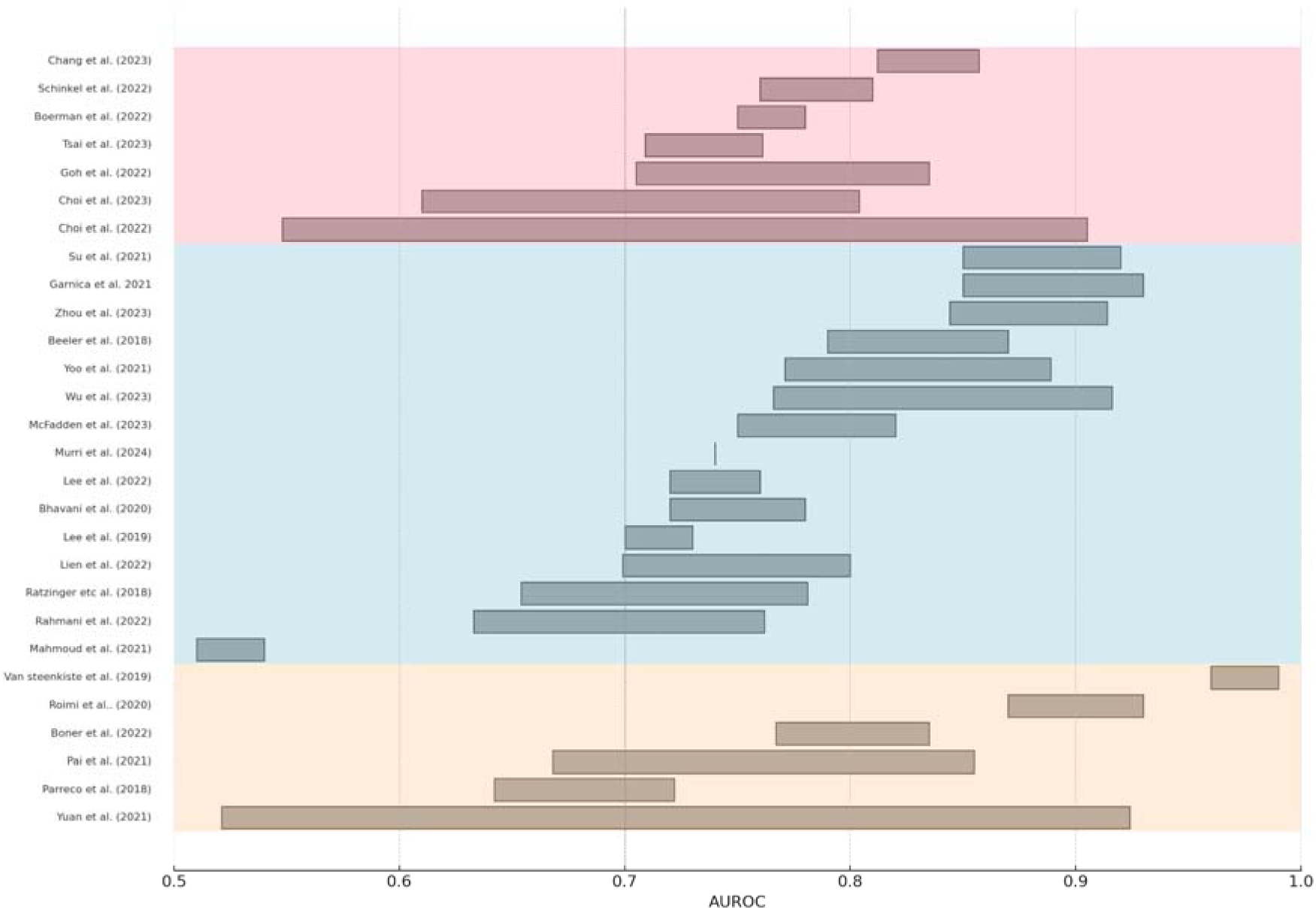
Horizontal bar chart depicting the spread of AUROC values reported in various studies. The studies are listed on the vertical axis, and each bar’s horizontal extension represents the range of minimum and maximum AUROC value achieved among the various models employed in the study. The color gradient red, indicates studies in ED, blue, indicating hospital inpatient settings, and the yellow area covers the studies in ICU settings.

**Fig 3.**
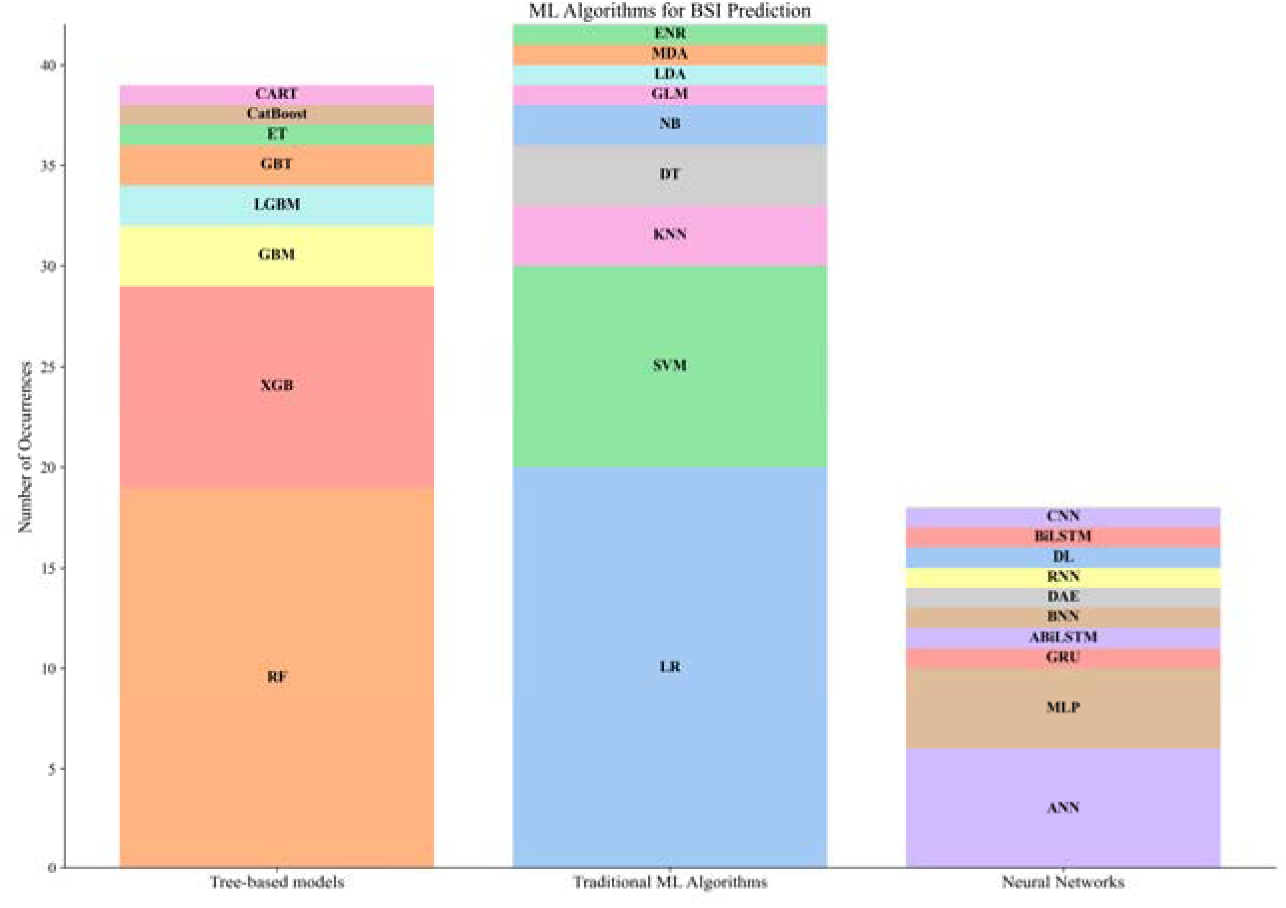
Stacked bar chart displaying the occurrence of MLalgorithms in BSI prediction studies across three categories: Tree-based models, Traditional ML Algorithms, and Neural Networks. For Tree-based models: Random Forest (RF), Extreme Gradient Boosting (XGB), Gradient Boosting Machine (GBM), Light Gradient Boosting Machine (LGBM), Extra Trees (ET), Gradient Boosted Trees (GBT), Classification and Regression Trees (CART), and Categorical Boosting (CatBoost). Traditional ML Algorithms include Logistic Regression (LR), Support Vector Machine (SVM), K-Nearest Neighbors (KNN), Decision Tree (DT), Naive Bayes (NB), Generalized Linear Model (GLM), Linear Discriminant Analysis (LDA), Multiple Discriminant Analysis (MDA), and Elastic Net Regression (ENR). Neural Networks are represented by Artificial Neural Network (ANN), Multilayer Perceptron (MLP), Bayesian Neural Network (BNN), Deep Learning (DL), Recurrent Neural Network (RNN), Gated Recurrent Unit (GRU), Convolutional Neural Network (CNN), Bidirectional Long Short-Term Memory (BiLSTM), Attention-based Bidirectional Long Short-Term Memory (ABiLSTM), and Denoising Autoencoder (DAE)

**Fig 4.**
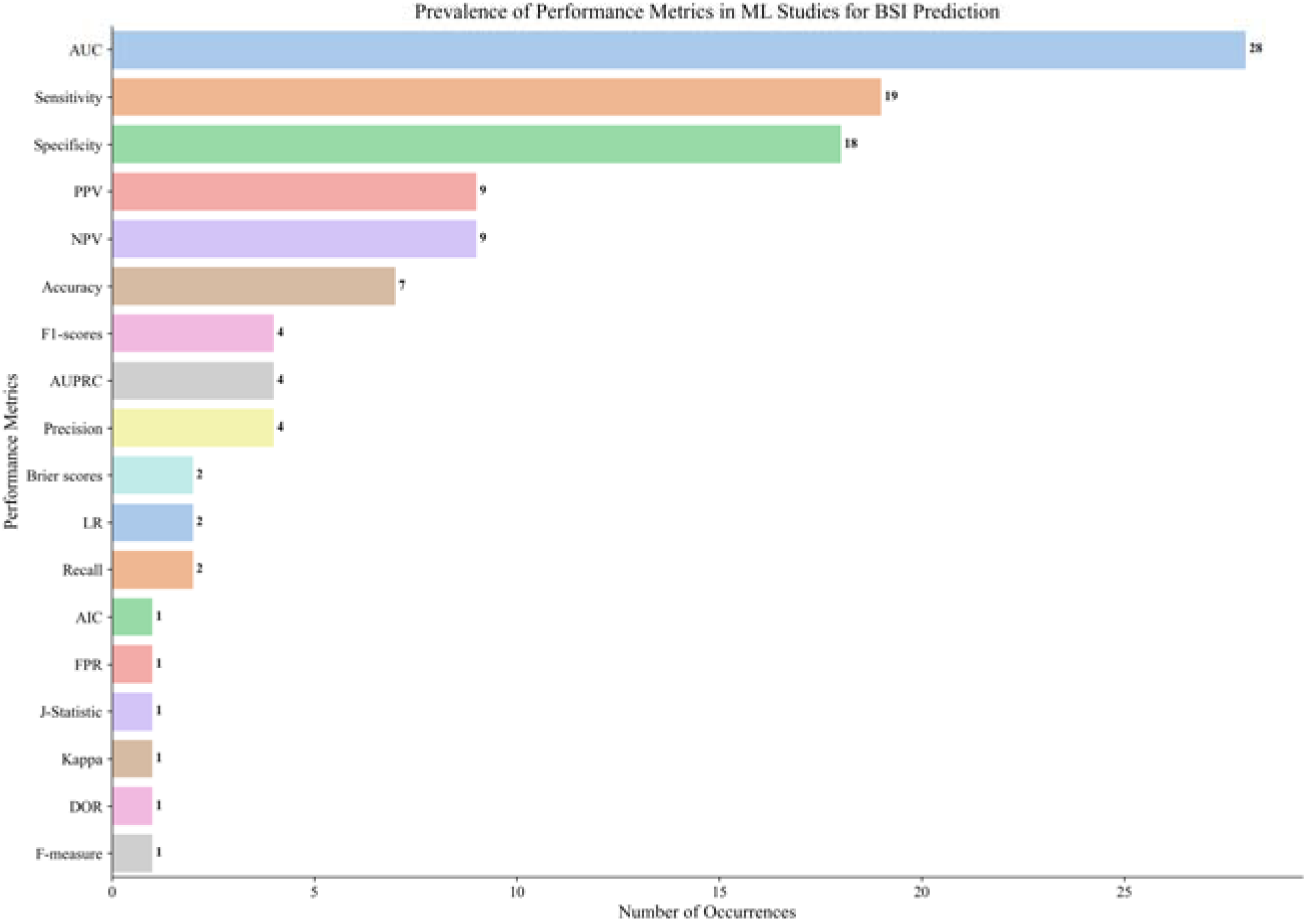
Horizontal bar graph summarizing the frequency of performance metrics reported in ML studies for BSI prediction. The metrics are displayed in descending order of occurrence, starting with Area Under the Receiver Operating Characteristic curve (AUC), followed by Sensitivity, Specificity, Positive Predictive Value (PPV), Negative Predictive Value (NPV), Accuracy, F1-scores, Area Under the Precision-Recall Curve (AUPRC), Precision, Brier scores, Likelihood Ratio (LR), Recall, Akaike Information Criterion (AIC), False Positive Rate (FPR), J-Statistic, Cohen’s Kappa, F-measure, and Diagnostic Odds Ratio (DOR)

**Fig 5.**
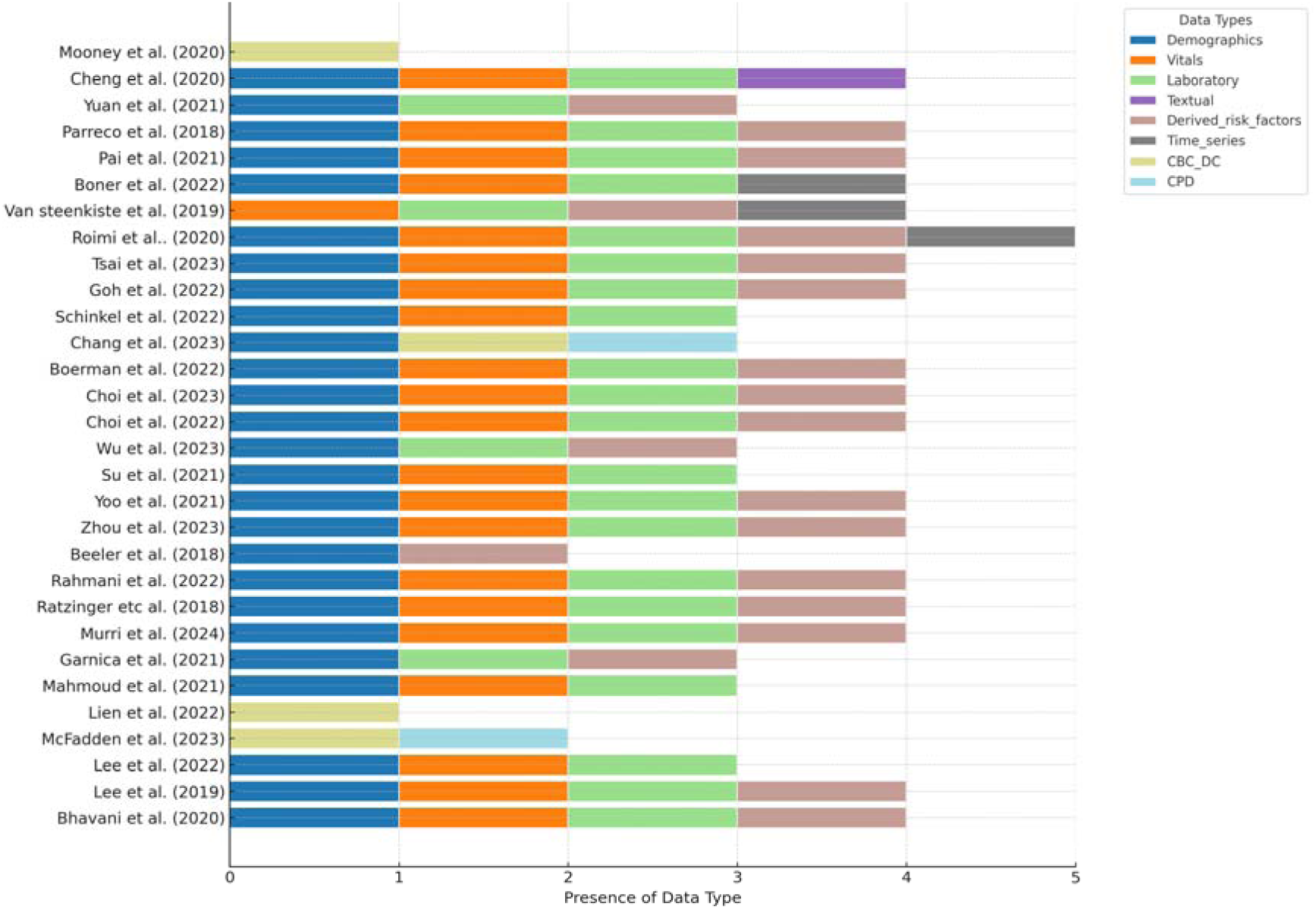
Stacked bar chart representing the types of data used in machine learning models for predicting bloodstream infections, across various studies listed on the vertical axis. Data types include demographics, vital signs, laboratory tests, textual data, derived risk factors, time-series data, Complete Blood Count/Differential Count (CBC/DC), and Cell Population Data (CPD). Each bar’s length indicates the number of data types used in each study, providing a comparison of data diversity across the research

**Table 2.** ML algorithms and key predictors in the selected studies.

| Setting | First author, year | ML models | Key predictors |
| --- | --- | --- | --- |
| Inpatients | Bhavani et al. | LR, GBM | Time from admission to BC, Temperature, Age, HR, Prior Bacteremia/Fungemia, |
|  | (2020) [20] |  | WBC, BUN, Glucose, DBP, SBP, PPI, RR |
|  | Lee et al. (2019) [21] | MLP, SVM, RF | ALP, PLT, Temperature, SBP, WBC, ICU stay, CRP, CVC, Age, PT, Hospital days to BC, HR, Gender, Antibiotics, RR, Creatinine |
|  | Lee et al. (2022) [22] | MLP, RF, XGB | PLT, Monocyte, Neutrophil, Bilirubin, Albumin, and Hospital stay, BUN, ALP, RR, PR, DBP, TP, WBC, PT, Hb, CRP, Creatinine, ALT, AST, Sodium, Chloride, ESR |
|  | Cheng et al. (2020) [23] | LR, NB, SVM, ADT, CNN, BiLSTM, ABiLSTM+ DAE | Textual chief complaints, Admission records, and Laboratory biochemical indicators. |
|  | McFadden et al. (2023) [24] | RF, XGB | CBC, DIFF, and CPD |
|  | Lien et al. (2022) [25] | RF, LR | CBC/DC, CRP, and PCT |
|  | Mahmoud et al. (2021) [26] | NN, RF, LR, DT, NB, SVM | Age, Antibiotics use, Surgery within 14 days, CVC, length of hospitalization before BC, RR, SBP, Temperature, DBP, HR, WBC, Sodium, PLT, Albumin, Creatinine, Lactic acid level. |
|  | Garnica et al. (2021) [27] | SVM, RF, KNN | The number of days in ICU before BC extraction, presence of Catheters, Chronic Respiratory disease, Fever, Age, CRP, PLT. |
|  | Murri et al. (2024) [28] | LR | Time BSI > 12 days, Procalcitonin > 1 ng/mL, Presence of a CVC, PLT, Hypotension, BUN, Presence of urinary catheter, Fever, Tachycardia, Altered mental status, Age, Bilirubin, Creatinine |
| Inpatients with SIRS | Ratzinger et al. (2018) [29] | RF, ANN, ENR | PCT, LBP, Albumin, Bilirubin |
| Inpatients with CVC | Rahmani et al. (2022) [30] | XGB, DT, LR | Temperature, HGB, comorbidities, Age, WBC, Race, Neutrophil. |
|  | Beeler et al. (2018) [31] | RF, LR | Age, Gender, history of CLABSI, CHG (Chlorhexidine Gluconate) Bathing Non-compliant Days, Line days. |
| HD patients | Zhou et al. (2023) [32] | LR, SVM, DT, RF, XGB | PCT, Temperature, Non-arteriovenous fistula dialysis access, NLR, Leukocyte, dialysis duration, LMR, Albumin, Neutrophil, PLT, Age, DBP, CRP, PLR, ALP, SBP, HR, history of BSI, |
| Cancer patients | Yoo et al. (2021) [33] | LR, ANN, RF, GBM, AML | Variables reflecting the dynamic status of patients with cancer, including blood urea nitrogen level, 7-day variance of RR, Total bilirubin level, 7-day variance of SBP, Body weight. |
| Maternity patients | Mooney et al. (2020) [34] | CART, LDA, KNN, SVM, RF | NLR, CBC parameters. |
| Patients with PCT $\leq 2.0$ ng/ml | Su et al. (2021) [35] | ANN, KNN, LR, RF, SVM, and NB. | Interleukin-6, PCT, D-dimer, Lactic acid, Leukocytes, Neutrophil, and PLT. |
| HIV patients | Wu et al. (2023) [36] | SVM, ANN, GBM, GLM, MDA, PLR, NB, RF | Low Hb, CD4+T cell, PLT, LDH, BUN, splenomegaly, absence of ART treatment, Strip shadow, Nodular shadow, and Shock. |
| ED | Choi et al. (2022) [37] | XGB, RF, LR | Chief complaint, Age, Temperature, HR, and DBP at triage stage. Neutrophils, PLT, CRP, Chief complaints, and Creatinine at disposition stage. |
|  | Choi et al. (2023) [38] | BNN | Age, HR, Temperature, DBP, History of chills, Ambulance use |
|  | Boerman et al. (2022) [39] | GBT, LR | Bilirubin, Urea, lymphocyte, Pulse rate, CRP, Neutrophil, age, Temperature, DBP, Potassium, Glucose, Thrombocytes, Creatinine, ALP, SBP, Organ damage |
|  | Chang et al. (2023) [40] | CatBoost, LGBM, XGB, RF, LR | Demographics, CPD, CBC/DC |
|  | Schinkel et al. (2022) [41] | XGB, LR | Temperature, Creatinine, CRP, Lymphocytes, DBP, Bilirubin, Thrombocytes, Neutrophils, ALP, HR, SBP, Leukocytes, Glucose, Age, Potassium, BUN, Sodium, monocytes |
| ED patients with SIRS | Goh et al. (2022) [42] | LR, SVM, RF | Age, Gender, COPD, Uncomplicated DM, Hemato-oncology, WBC, Band cell, Platelet, Temperature, HR, mild liver disease, Mean arterial pressure, RR, GSC |
| ED patients with fever | Tsai et al. (2023) [43] | RF, LR, MLP, XGB, LGBM | Hypertension, Gender, Temperature, DM, Age, CRP, PLT, WBC, Malignancy, Eosinophil, HR, BMI, Hb, RR, SBP, DBP, Band, CKD, Liver Cirrhosis, COPD, GCS |
| ICU patients | Roimi et al. (2020) [44] | RF, XGB | Time duration (days) between sampling time and last defecation, Time duration (hours) between sampling time and the maximum BUN (mg/dL) value measured during the 5 days prior to sampling, Length of stay (days) between sampling time and ICU admission, The minimal weight (kg) during the 5 days prior to sampling, The time duration between sampling time and the maximum MCHC (g/dL) during the 5 days prior to sampling |
|  | Van Steenkiste et al. (2019) [45] | BiLSTM, ANN, SVM, KNN, LR | Temperature, Thrombocytes, Leukocytes, CRP, sepsis-related organ failure assessment, HR, RR, PT, and mean systemic arterial pressure. |
|  | Boner et al. (2022) [46] | FNN, GRU, CNN, LR | Temperature, BUN, BP, HR, Albumin, PLT, Chloride, Creatinine, Chloride, and Phosphorus. |
|  | Pai et al. (2021) [47] | LR, SVM, MLP, RF, XGB | ALP, CVC period, prothrombin time, PLT, Albumin, Apache II score, Age, foley |
| ICU patients with CVC | Parreco et al. (2018) [48] | LR, GBT, DL | Severity of illness scores (like SAPS II, APS III, and OASIS) and comorbidities. |
| ICU patients with new-onset SIRS | Yuan et al. (2021) [49] | XGB, SVM, RF, ET, LR | Colonization, Diabetes, AKI, total number of parenteral nutrition days, history of fungal infection, CRRT days, Abdominal surgery, BDG, days of mechanical ventilation, Length of hospital and ICU stay, days of CVC |

This table summarizes the ML algorithms and key predictors from the selected studies

### 3.4. Data challenges and strategies

BSIs may be relatively rare compared to the number of non-infection cases in a dataset. This imbalance can lead models to become biased towards predicting the majority class, reducing their effectiveness in identifying true infection cases. Most studies reported imbalanced dataset with prevalence rates of BSI as given in the Table 1. To overcome challenges with data imbalance, Synthetic Minority Over-sampling Technique (SMOTE) was widely used for augmenting the minority class in the dataset by generating synthetic samples [21, 30, 49, 51]. Goh et al. (2022) employed oversampling, undersampling, and random oversampling (ROSE) methods for model development [42]. Lien et al. (2022) and Van Steenkiste et al. (2019) employed Precision-Recall Area Under Curve (PRAUC) metric for a more accurate assessment of model performance in imbalanced datasets [25, 45].. The study by Garnica et al. (2021) encountered significant issues with missing data across the patient records used [27]. The types of missing data were classified into three categories: Missing Completely At Random (MCAR), Missing At Random (MAR), and Missing Not At Random (MNAR) [52]. They employed separate class method to represent the missing data, ensuring the ML models could handle these cases without dropping significant amounts of data. Using patient data for training ML models can raise concerns about privacy and data security. Ensuring patient anonymity and complying with regulations can limit the accessibility and use of certain data. Boerman et al. (2022) faced difficulty with the limitation of not being able to use free-text data such as physician and nurse reports due to privacy concerns [39]. To ensure patient privacy and compliance with data protection regulations researchers can implement effective deidentification of patient records, involving elimination or alteration of direct identifiers, such as names, age, gender, or location, which could be combined to identify an individual.

### 3.5. Quality of evidence and risk of bias

The risk of bias for the retrospective diagnostic test accuracy studies was assessed following the QUADAS-2 guidelines, and the results are shown in Table 3. For patient selection, six articles (20.6%) presented high risk of bias and four articles (13.7%) presented unclear risk of bias for failing to describe the study population and patient selection. For index test 12 articles scored a high risk of bias (41.3%) and five article (17.2%) scored unclear risk of bias, due to non-reporting of data splits and cross-validation strategies. Three articles (10.3%) scored high risk of bias and one score unclear risk of bias for not specifying the reference standards. One article presented unclear risk of bias in the flow and timing.

**Table 3.**
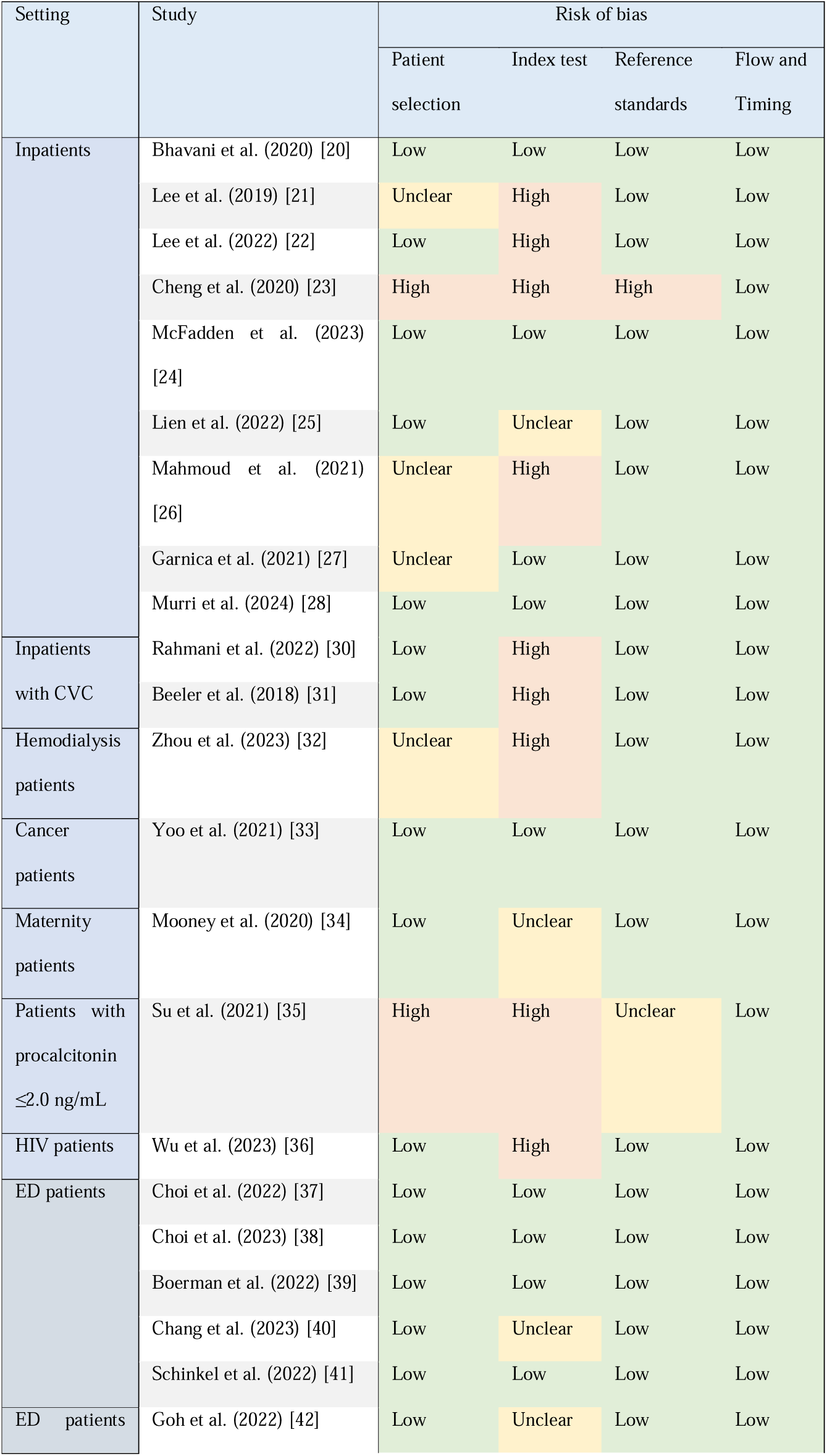
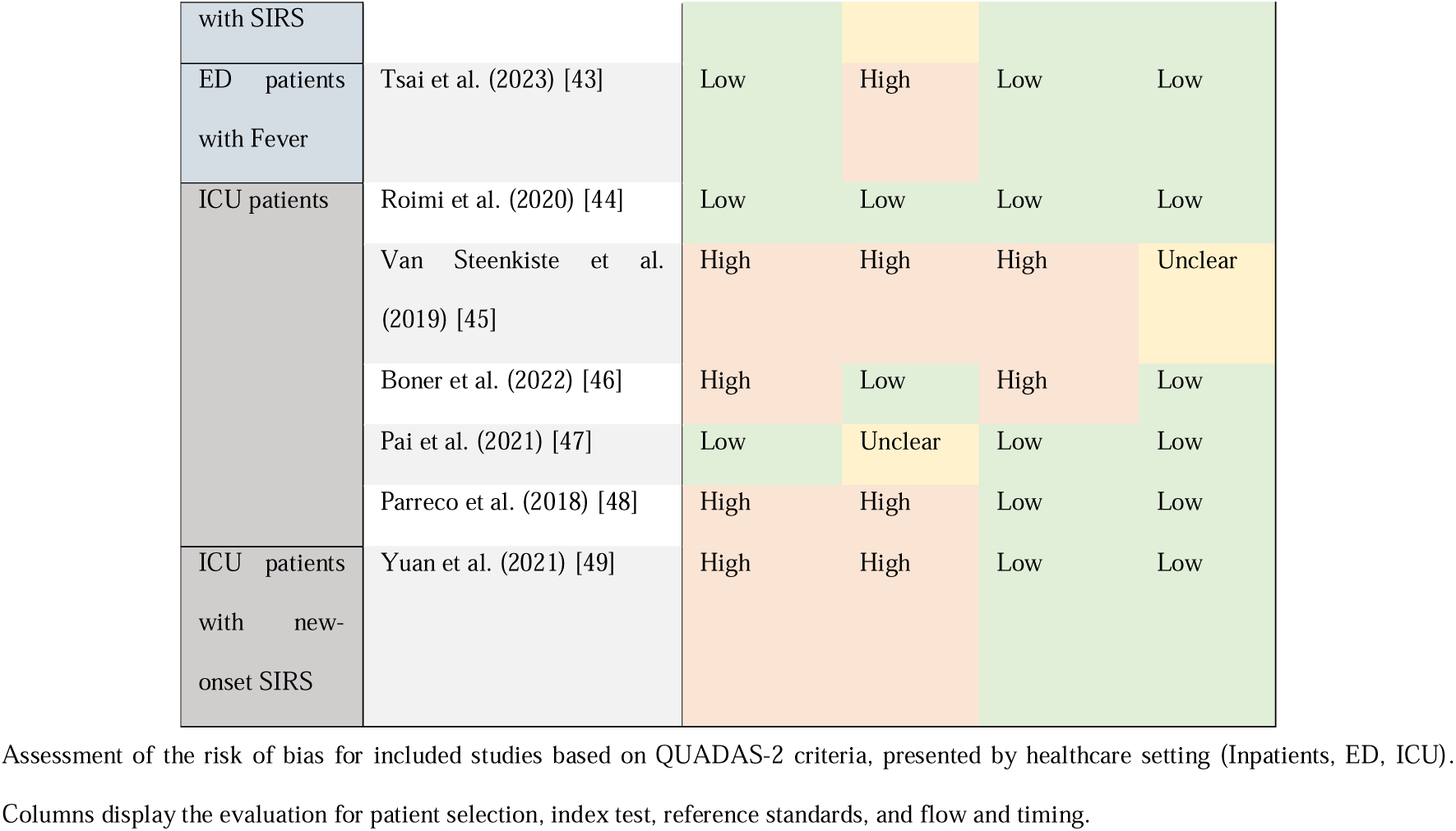
Risk of Bias Assessment Results According to QUADAS-2 Criteria.

The GRADE evidence profile was estimated by pooling studies based on settings. In this analysis we only considered studies focusing on general adult patient population and did not consider studies reporting specific study population. The GRADE evidence profile is given as Table 4. The studies in ICU and Inpatients settings aggregates were considered at high risk of bias and studies in ED settings aggregates were considered unclear risk of bias. One level of evidence was deducted for observational study design and Inconsistencies due to heterogeneity. Consequently, the quality of evidence for each of the settings was scored as low. Additionally, the outcome column distinguishes AUROC values for high and unclear risk of bias studies. High risk of bias studies reported lowest AUROC values in the inpatient settings and reported highest AUROC values in the ED and ICU settings.

**Table 4.** GRADE Evidence Profile and Quality Assessment of Studies by Setting.

| Settings | Studies Characteristics |  | Quality Assessment |  | Outcome |  |
| --- | --- | --- | --- | --- | --- | --- |
|  | Number of studies | Design | Limitations (Unclear risk of bias studies/total) | Inconsistency | AUROC high risk of bias/unclear risk of bias | Quality of evidence |
| Inpatients | 9 studies [20-28]<br>(1,195,629)<sup>a</sup> | Observational cohort studies | High risk of bias (5/8) | High heterogeneity | 0.51-0.762/ 0.74-0.86 | Low |
| ED | 5 studies (72,072)<br>[37-41] | Observational<br>cohort studies | Unclear risk of bias<br>(1/5) | Moderate<br>heterogeneity | 0.834-0.844/ 0.728-0.782 | Low |
| ICU | 4 studies (16,381)<br>[44-47] | Observational<br>cohort studies | High risk of bias (2/4) | Considerable<br>heterogeneity | 0.835-0.97/ 0.668-0.851 | Low |

This table shows the GRADE evidence profile and overall quality of studies segregated by hospital setting (Inpatient, ED, ICU).
Note: <sup>a</sup> 4 studies in this pool did not report number of patients and so the reported number of BSI episodes were included to calculate the sum.

### 3.6. Meta-analysis

In the meta-analysis, only models presented in articles with target condition bacteremia and general patients population across the three settings were considered. Models for specific disease population, models not reporting AUROC metric, and models predicting CLABSIs were not considered in the meta-analysis. A total of 41 models and 9 covariates were included in the meta-analysis. The univariate and multivariate random effect model significant (p-value>0.05) results are shown in Table 5. The random effect model analysis shows that laboratory tests, use of time-series data, use of CBC/DC data, BiLSTM model, XGB model, and RF model positively contributed to the AUROC. We performed a pooled analysis by setting, to identify the best covariates for each. The results of the regression analysis are given as Table 6. For Inpatient setting, demographics (includes age), laboratory tests, and use of CBC/DC data positively contributed to the model performance and use of vital signs negatively impacted the model performance. In contrast in the ICU setting, vital signs, laboratory tests, derived risk factors, use of time-series data, and BiLSTM model employed for sequential data positively impacted the performance metrics.

**Table 5.**
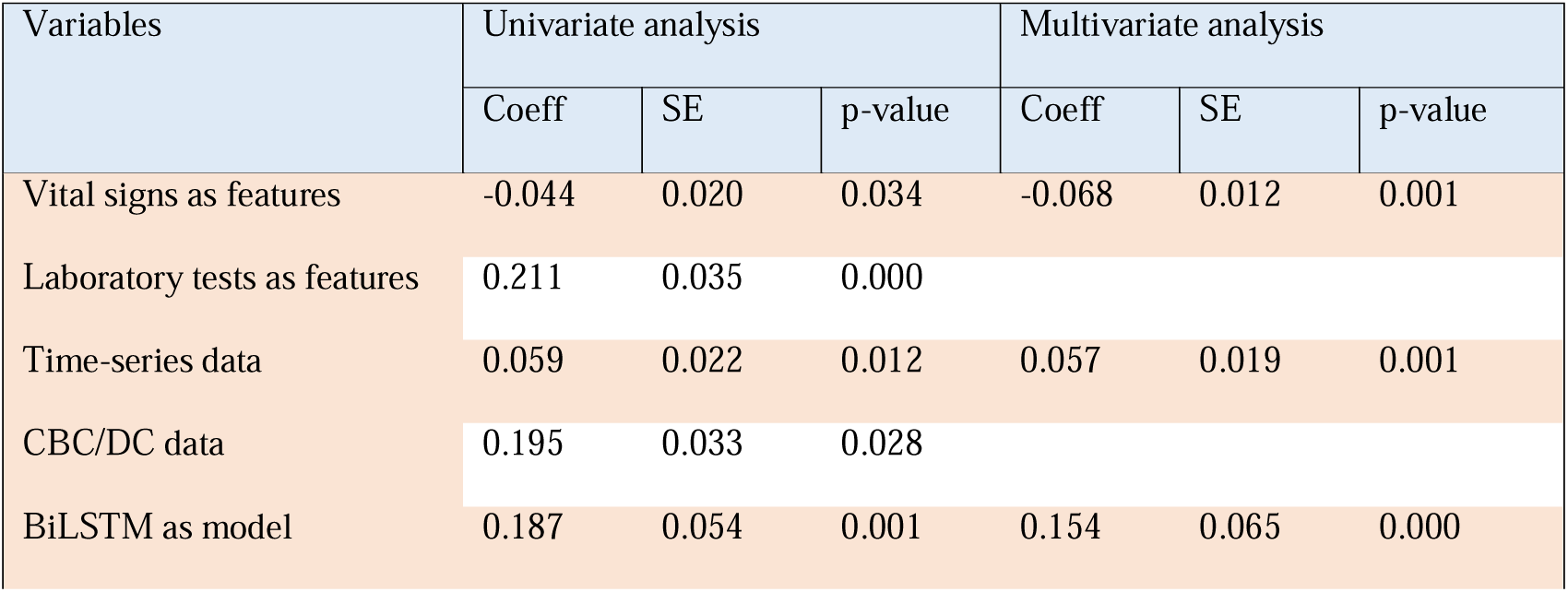
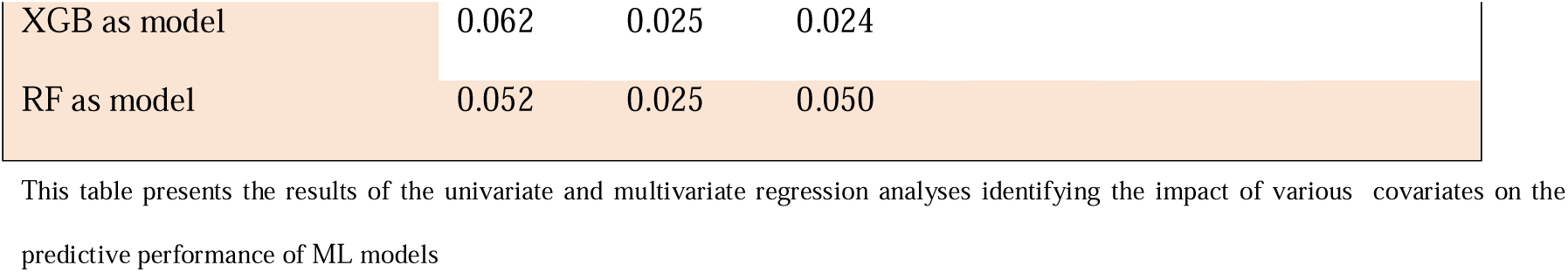
Univariate and Multivariate Analysis of Predictive Performance Covariates.

| Variables | Univariate analysis |  |  | Multivariate analysis |  |  |
| --- | --- | --- | --- | --- | --- | --- |
|  | Coeff | SE | p-value | Coeff | SE | p-value |
| Vital signs as features | -0.044 | 0.020 | 0.034 | -0.068 | 0.012 | 0.001 |
| Laboratory tests as features | 0.211 | 0.035 | 0.000 |  |  |  |
| Time-series data | 0.059 | 0.022 | 0.012 | 0.057 | 0.019 | 0.001 |
| CBC/DC data | 0.195 | 0.033 | 0.028 |  |  |  |
| BiLSTM as model | 0.187 | 0.054 | 0.001 | 0.154 | 0.065 | 0.000 |

|  |  |  |  |
| --- | --- | --- | --- |
| XGB as model | 0.062 | 0.025 | 0.024 |
| RF as model | 0.052 | 0.025 | 0.050 |

This table presents the results of the univariate and multivariate regression analyses identifying the impact of various covariates on the predictive performance of ML models

**Table 6.** Pooled Regression Analysis Results by Study Setting.

| Variables | Regression analysis |  |  |
| --- | --- | --- | --- |
|  | Coeff | SE | p-value |
| **Inpatients settings:** |  |  |  |
| Demographics | 0.184 | 0.010 | 0.000 |
| Vitals as features | -0.138 | 0.010 | 0.001 |
| Laboratory features | 0.184 | 0.010 | 0.000 |
| CBC/DC data | 0.282 | 0.006 | 0.000 |
| **ED settings:** |  |  |  |
| None with significant p-value |  |  |  |
| **ICU settings:** |  |  |  |
| Vitals as features | 0.131 | 0.009 | 0.005 |
| Laboratory features | 0.131 | 0.009 | 0.005 |
| Derived risk factors as features | 0.184 | 0.026 | 0.020 |
| Time-series data | 0.100 | 0.024 | 0.050 |
| BiLSTM as model | 0.250 | 0.033 | 0.017 |

This table presents the coefficients, standard errors, and p-values from the regression analysis of significant covariates affecting model performance in different hospital settings (Inpatient, ED, ICU).

## 4. DISCUSSIONS

This is the first systematic review and meta-analysis that has integrated findings from diverse clinical settings to assess the performance of ML models in predicting BSIs. This work corroborates the growing evidence that ML models are viable tools for enhancing diagnostic accuracy and potentially reducing reliance on traditional blood cultures, a point underscored by contemporary studies [3, 6]. Significantly, our meta-analysis revealed that models performed variably across different clinical environments, with AUROC scores ranging widely. In the inpatient settings, AUROCs ranged from 0.51 to 0.866, demonstrating moderate to high diagnostic accuracy. This performance variance was even more pronounced in the ICU settings, where AUROCs spanned from 0.668 to 0.970, suggesting that certain models are highly effective while others may require further refinement. The variability in model performance aligns with the findings from previous studies which have highlighted the complexity involved in creating universally effective ML models for BSI prediction [16]. This analysis not only validates the potential of ML in clinical diagnostics but also highlights the critical need for tailored approaches depending on specific hospital settings and patient populations. Such differentiation in model efficacy emphasizes the importance of context-specific model development and deployment, which should be informed by the distinct dynamics and needs of each clinical environment. Furthermore, the integration of various data types, such as demographic features, laboratory tests, and derived clinical features, has been shown to significantly impact model performance, echoing the findings of recent meta-analyses which advocate for a multi-faceted approach to data integration within ML models to enhance diagnostic precision and reliability [53].

### 4.1. Clinical relevance and model performance

The findings from our study underscore the potential of ML models to enhance the diagnostic accuracy of bloodstream infections BSIs significantly. This enhancement is crucial, considering the current reliance on traditional blood cultures, which, while standard, are not without their limitations such as delays in results and the potential for contamination [54]. ML models, by leveraging a variety of clinical data, including patient demographics, prior medical history, and laboratory results, can predict BSIs with notable accuracy. This predictive capability is especially valuable in clinical settings where rapid decision-making is critical [55]. By predicting BSIs accurately, ML models can facilitate earlier intervention strategies, potentially leading to improved patient outcomes and reduced mortality rates [56]. Furthermore, the integration of ML into hospital systems offers a pathway to more streamlined resource allocation [57–59]. By accurately identifying patients at high risk of BSIs, hospitals can optimize the use of tests and allocate personnel and medical resources more efficiently [60]. This not only helps in managing hospital resources but also in reducing unnecessary antibiotic use, which is often a knee-jerk response to suspected infections [61].

### 4.2. Future directions and academic contribution

Our study have laid the groundwork for several promising directions that future investigations could take to advance this critical area of medical informatics. While retrospective studies form the bulk of current research on ML models for BSI prediction, there is a pronounced need for prospective studies. Such studies will allow for real-time data collection and model validation, offering insights that are often obscured in retrospective analyses [12]. This shift could also facilitate the adjustment of models in accordance with dynamic clinical environments, ensuring that they remain robust and reliable over time. The potential of ML models to be integrated into real-time clinical decision support systems represents a significant leap towards operationalizing AI in everyday clinical practice [62]. However, achieving this requires rigorous testing and validation of these models within clinical settings to ensure they perform reliably when interfaced with live data streams [63, 64]. There is substantial scope for exploring new predictive variables that could enhance the predictive accuracy of ML models. The incorporation of genomic data, patient mobility patterns, and real-time monitoring data could provide new insights into infection risk factors, potentially leading to more sophisticated and accurate prediction models [65, 66]. To build trust and validate the efficacy of ML models in clinical settings, systematic reporting and external validation are essential [67, 68]. Models need to be tested across diverse demographics and varied clinical environments to assess their universality and reliability [69–72]. The work of Fleuren et al. (2020) and Moor et al. (2021) highlights the importance of such validation in confirming the utility and accuracy of predictive models for sepsis, which can be paralleled in BSI prediction [12, 13]. As ML applications in healthcare continue to expand, it is crucial to consider the regulatory and ethical implications of their use [73, 74]. Ensuring patient privacy, securing data, and maintaining transparency in AI decision-making processes are paramount [75, 76]. Future research must also address these aspects to foster a safe and trustful adoption of AI technologies in healthcare.

### 4.2. Strengths and Limitations

Our systematic review and meta-analysis are grounded in a comprehensive, methodologically robust approach that integrates a variety of data sources and analytical techniques. The extensive database search across multiple platforms including PubMed, IEEE Xplore, and Scopus ensures a broad capture of relevant studies, reducing the risk of publication bias. This wide-ranging data acquisition is supplemented by our application of the QUADAS-2 framework and GRADE methodology, which enhances the credibility of our findings by systematically assessing the risk of bias and the quality of evidence across studies. Furthermore, the synthesis of data into figures and comprehensive tables enables clear visual representation and understanding of the applications of ML models across varied clinical settings. Our findings are supported by rigorous statistical analysis, including univariate and multivariate models, which reveal key performance drivers and validate the predictive power of ML models for bloodstream infections. Despite these strengths, several limitations must be acknowledged. A significant proportion of the included studies utilized retrospective designs. While retrospective studies provide valuable historical insights, they are inherently limited by the data available, often lacking the prospective validation needed to confirm the efficacy of predictive models under current clinical conditions. This design limitation impacts the generalizability of our findings, as the models might perform differently when deployed in real-time environments. To enhance the generalizability and applicability of future research, several strategies can be adopted. Prospective study designs should be prioritized, as they allow for the real-time evaluation and adjustment of ML models, ensuring that the models are tested and validated under varied clinical conditions. This approach not only tests the robustness of the models but also helps in fine-tuning them for practical deployment. Furthermore, multi-center studies involving diverse populations and settings should be encouraged to test the efficacy of these predictive models across different demographics and operational conditions. Such studies can help identify and mitigate any population-specific biases, thereby enhancing the models’ applicability and reliability. Lastly, ongoing validation and systematic reporting should be integral to future research efforts. By continuously assessing and reporting the performance of ML models, researchers can ensure that these tools remain effective and relevant in the ever-evolving clinical landscape. By addressing these limitations and building on the strengths of our current approach, future research can significantly advance the field of ML in medical diagnostics, particularly in the prediction and management of bloodstream infections.

## 5. CONCLUSIONS

Our systematic review and meta-analysis have critically evaluated the efficacy of ML models in predicting BSIs, a crucial domain where timely and accurate diagnosis can significantly influence clinical outcomes. ML models, especially those incorporating diverse data types such as laboratory results and demographic information, demonstrated a capacity to predict BSIs with a high degree of accuracy. The integration of these models into clinical settings can potentially reduce the reliance on traditional blood cultures, which, while foundational, are hampered by delays and susceptibility to contamination. This shift could streamline diagnostic workflows and enhance the speed and precision of infection response interventions, thus improving patient care and outcomes. To advance the use of ML in predicting BSIs, future research should focus on prospective studies and the development of real-time, adaptive ML systems that can be integrated seamlessly into clinical decision-support frameworks. Furthermore, external validation of these models across diverse populations and settings is essential to bolster clinical confidence and foster wider adoption. In conclusion, while ML models hold significant promise for transforming BSI diagnosis, their successful implementation will depend on meticulous model development, validation, and customization tailored to the nuanced demands of different healthcare settings. This holistic approach will be crucial in overcoming the current challenges and fully realizing the potential of ML in clinical diagnostics.

## Data Availability

All data produced in the present work are contained in the manuscript

## Acknowledgements

We would like to thank the researchers at Mid-Norway Centre for Sepsis Research for valuable discussions and feedback.

## Author’s contributions

RB conceptualized and designed the study with inputs from JKD and ØN. RB performed database search and literature analysis. RB wrote the initial and subsequent drafts of the manuscript, which other authors reviewed and approved.

## Appendix List of Medical Abbreviations

- ALP: Alkaline Phosphatase
- ALT: Alanine Aminotransferase
- APS III: Acute Physiology Score III
- ART: Antiretroviral Therapy
- AST: Aspartate Aminotransferase
- AKI: Acute Kidney Injury
- BC: Blood Cultures
- BDG: Beta-D-Glucan
- BUN: Blood Urea Nitrogen
- CBC: Complete Blood Count
- CHG: Chlorhexidine Gluconate
- CKD: Chronic Kidney Disease
- CLABSI: Central Line-Associated Bloodstream Infection
- COPD: Chronic Obstructive Pulmonary Disease
- CPD: Cephalopelvic Disproportion
- CRP: C-reactive Protein
- CRRT: Continuous Renal Replacement Therapy
- CVC: Central Venous Catheter
- DBP: Diastolic Blood Pressure
- DIFF: Differential Count
- DM: Diabetes Mellitus
- ESR: Erythrocyte Sedimentation Rate
- GCS: Glasgow Coma Scale
- Hb: Hemoglobin
- HR: Heart Rate
- ICU: Intensive Care Unit
- LMR: Lymphocyte to Monocyte Ratio
- NLR: Neutrophil to Lymphocyte Ratio
- OASIS: Oxford Acute Severity of Illness Score
- PCT: Procalcitonin
- PLR: Platelet to Lymphocyte Ratio
- PLT: Platelet Count
- PPI: Proton Pump Inhibitor
- PT: Prothrombin Time
- RR: Respiratory Rate
- SAPS II: Simplified Acute Physiology Score II
- SBP: Systolic Blood Pressure
- WBC: White Blood Cell Count

